# Prior antiretroviral therapy exposure among clients presenting for HIV treatment initiation in South Africa: an exploratory mixed-methods study using multiple indicators of exposure

**DOI:** 10.1101/2024.08.23.24312454

**Authors:** M Benade, M Maskew, V Ntjikelane, N Scott, N Ngcobo, BE Nichols, L Malala, M Manganye, S Rosen

## Abstract

**Background:** The era of universal treatment for HIV has seen high rates of disengagement from antiretroviral therapy (ART) programs and re-engagement after interruptions, with modeled estimates of non-naïve initiators >50% in many places. Most re-engagers are reluctant to admit prior antiretroviral exposure, and non-self-reported data on proportions of re-initiators are scarce. We synthesized data from multiple sources to explore the proportion of people who present for initiation with evidence of prior ART use in South Africa.

**Methods:** We enrolled a sequential sample of adults presenting to initiate ART or to re-initiate ART after an interruption >3 months and collected 1) self-reported previous treatment experience; 2) electronic medical record (EMR) evidence of prior ART clinic visits; 3) baseline blood tests for metabolites of tenofovir diphosphate; and 4) laboratory records indicating prior ART-related tests. Interviews were conducted with a sub-sample of clients who self-reported no prior ART use but had evidence of metabolites.

**Results:** Among 89 enrolled participants (median age 32.5, 62% female), 16 (18%) self-reported previously taking ART >3 months prior to enrolment. An additional 33 (45%) who did not self-report prior exposure had EMR or laboratory evidence of prior ART use, for a total of 49 (55%) clients with known prior treatment exposure at initiation. Sensitivity of self-report was 40%, EMR 43%, metabolite testing 45%, and laboratory records 73%. Interviewees (n=11) reported opting to present as naive because they perceived that disclosure of prior disengagement would cause delays accessing treatment, require additional documentation, and elicit negative responses from healthcare workers. Study limitations included short duration of metabolite detectability (90 days), inability to link individuals within the EMR to discern ART experience at other facilities, and lack of baseline viral load testing.

**Conclusions:** At least 55% of clients initiating ART in South Africa have prior treatment experience, but only a third of re-initiators voluntarily reveal this. Laboratory records, which reflect long-term experience, yielded the most accurate results for ascertaining prior treatment exposure. As numbers re-engaging in HIV care after a treatment interruption increase, understanding reluctance to self-report ART experience and exploring opportunities to overcome barriers are critical for preventing repeated interruptions.

## Introduction

As rates of HIV treatment with antiretroviral therapy (ART) coverage approach the global target of 95% of HIV-positive individuals who are aware of their status[1], national HIV programs in sub-Saharan Africa are experiencing very high rates of both disengagement from treatment and re-engagement after interruptions[2,3]. As a result, a large proportion of clients who present at healthcare facilities for treatment initiation are not ART-naïve--they have prior ART exposure and are instead re-starting treatment after a short or long interruption. We recently conducted a systematic review of studies reporting re-initiation proportions and found estimates ranging from a low of 2% of initiators to a high of 53%[4]. Since then, researchers in South Africa’s Western Cape Province have reported re-initiation rates as high as 69%[3].

The take-home message of our review[4] was that data on the proportion of clients who are re-initiators are scarce. The review found only 11 primary data estimates for all of sub-Saharan Africa. The review identified four indicators of prior exposure. Many of the estimates were based on 1) client self-report, and others on 2) baseline (presentation for ART initiation) viral load test results showing existing viral suppression. Others located 3) earlier viral load test results in clients’ clinic or laboratory records, indicating a previous period of treatment. Finally, some studies utilized laboratory testing to seek 4) ARV metabolites in blood, hair, or urine samples. Each of these indicators is constrained in some important way. Self-report, which produces the lowest estimates, suffers from clients’ reluctance to reveal previous disengagement from care. Both viral load suppression at baseline and metabolite tests capture only recent interrupters, while medical records, which have the potential to reveal much earlier ART use, often cannot be located, cannot be linked to the same individual’s clinic record, or are incomplete[5]. Adding to the difficulty in interpreting results is that many of the studies including in the review that did not rely on self-report explicitly excluded anyone who did self-report prior exposure.

Clients who present for treatment initiation with prior ART exposure have, by definition, disengaged from or interrupted treatment in the past. If the reason for the prior interruption has not been addressed, these clients may face greater or different obstacles to long-term retention in care than do naïve patients. Understanding the proportion of initiators who are non-naïve and being able to match them with suitable interventions are thus important for improving treatment program outcomes. To start to build a reliable evidence base on this topic for sub-Saharan Africa, we conducted an exploratory, mixed-methods sub-study that collected multiple indicators of prior use for a sample of ART initiators in South Africa.

## Methods

### Study sites and population

The study sequentially enrolled adults (≥18) who reported either newly initiating ART or re-initiating ART after an interruption of more than 90 days at one of three public sector primary healthcare facilities in South Africa, one each in Mpumalanga, KwaZulu-Natal, and Gauteng provinces. The study sites are a subset of facilities participating in the AMBIT SENTINEL study described elsewhere [6]. Participants enrolled in this study were also enrolled in the PREFER survey[7], a questionnaire-based study of preferences for services in a patient’s first six months on antiretroviral therapy for HIV, conducted at the same facilities as the SENTINEL study.

For this sub-study of prior ART experience, a sequential subsample of PREFER participants who self-reported that they were initiating ART for the first time or re-initiating ART after an interruption of at least 90 days were asked for additional consent to create a dried blood specimen (DBS) from their routinely collected blood samples and to have the DBS tested for the presence of ARV metabolites, as described below. Most participants who consented to DBS creation completed the full PREFER survey, although a small number were administered an abbreviated instrument that asked only about prior treatment experience, not the full set of preferences and other questions. Eligibility for the sub-study was confirmed using a survey screening form. PREFER participants who consented and completed study procedures received a voucher equivalent to USD10 as a token of thanks for their time and participation.

Sample size for the sub-study was based on time and resource availability, rather than any expectation of findings. Delays due to ethics approvals, and study site willingness to create dried blood spot samples in a smaller sample than originally hoped[7], but enough samples were collected to provide exploratory results.

### Data collection

Data were collected from five sources, as described in Table 1.

**Table 1.** Sources of data and corresponding outcome measures for the sub-study.

| Source | Description | Outcome measured | Strengths and weaknesses of source |
| --- | --- | --- | --- |
| Survey | The PREFER questionnaire was administered to participants by a trained research assistant upon presentation for ART initiation or re-initiation and provision of written consent. PREFER asked questions about prior experiences of HIV care and treatment, barriers to retention in care, and clients' preferences and expectations for receiving care. The consent form for PREFER allowed access to participants' medical records and the blood specimens routinely collected by clinic staff at treatment initiation. | "Self-reported ART use" as reported during the survey. Responses included 1) No previous reported ART use; or 2) Previous ART use, but no ART use in the 90 days prior to ART initiation. (Participants with prior ART use in the preceding 90 days were deemed ineligible for this substudy at screening.) | Strengths: No time limitation--self-report should capture any ART use from first exposure.<br>Weaknesses: Disclosure is rare; most ART clients do not reveal prior ART experience. |
| Blood samples for metabolite testing | Nurses at the study sites were trained to prepare dried blood specimen (DBS) samples from the blood specimens they collected as part of routine care at ART initiation. All DBS samples were created on the same day that the participant's venous blood sample was collected as part of routine care at ART initiation[8] and stored in a clinic freezer prior to shipping to the analyzing laboratory. Specimens were shipped by air freight on dried ice with temperature monitoring to the Division of Clinical Pharmacology at the University of Cape Town, South Africa. Mass spectrometry was performed to measure for metabolites of tenofovir diphosphate (TDF), which are typically detectable for approximately 90 days, allowing detection of prior use for up to 3 months [9]. Because patients who self-reported any ART use $\leq 90$ days prior to study screening were excluded from the study, all metabolite tests would be negative if self-disclosure were accurate. Further details of DBS collection are outlined in Supplementary file 1. | We created a dummy variable labeled "evidence of recent ART use by DBS specimen", where any detectable TDF was deemed positive. Those with evidence were classified as having levels consistent with regularly taking 4-6 doses per week, 2-3 doses per week, or <2 doses per week. | Strengths: Quantified concentration data from biological sample.<br>Weaknesses: Short term; metabolites become undetectable after approximately 90 days, meaning that ART use >90 days prior to presentation would not be detectable. |
| Electronic medical records | South Africa's electronic medical record for HIV treatment, TIER.Net[5], was accessed for study patients to search for prior HIV treatment interactions with the healthcare system, including clinical consultations and prescriptions. We note that this data source is expected to be quite incomplete over time, as it does not use a consistent identification number to distinguish individual patients. A patient presenting at a different facility, or even at the same facility after an interruption, may be assigned a new TIER.Net record number, effectively concealing any previous receipt of services. | Any entry in an individual's EMR record since the earliest date in the dataset (2013) indicating ART initiation, monitoring, or medication prescribing or dispensing was designated as "evidence of previous ART use by EMR." | Strengths: No time limitation.<br>Weaknesses: No unique patient identifier; misses prior initiations of the same client if at a different healthcare facility. |
| Laboratory records | In South Africa, samples collected for laboratory testing by the National Health Laboratory Service (NHLS) are identified with a unique specimen number (barcode) at the collection facility. We recorded these numbers for the routine blood samples collected on day of ART initiation and reviewed results in the NHLS online portal to see if participants had any evidence of prior ART use, such as any prior viral load test results. | "Evidence of previous ART use by lab," defined as having had one or more viral load tests in the past. | Strengths: No time limitation; can capture prior treatment experience at other facilities; more sensitive than EMR data when using date of birth and name to search.<br>Weaknesses: Viral load testing for monitoring purposes is typically only done after six months in care, therefore underestimating prior exposure for those who initiated care but did not remain in care long enough to qualify for viral load testing. |
| Qualitative interviews | A subset of participants who self-reported no prior ART use but were found to have ARV metabolites in their blood samples were contacted for telephonic interviews to understand their reasons for not disclosing prior exposure [10]. Additional questions included how they felt on the day of the interview, what their circumstances were, what staff could have done to make it easier to disclose, and why they think others may not disclose prior treatment history. All participants in these interviews provided written informed consent to be contacted. Interviews lasted approximately 15 minutes, were audio recorded, translated into English, and transcribed verbatim for analysis. | Qualitative findings from interviews, key emerging themes. | Strengths: Contextual understanding, the interviews provided in-depth insights into why participants opted to present as naive prior to their exposure to antiretroviral therapy.<br>Weaknesses: Due to the small, non-random sample used in this qualitative subset, the findings from these interviews may not be broadly generalizable to larger populations. |

Another indicator of prior ART exposure is viral suppression at baseline (initiation). Routine, baseline viral load testing is not recommended in South African guidelines, however, and only a small number of participants in the sub-study did receive viral load tests from the clinic when they presented for initiation. This was done at the discretion of clinic staff, most likely because the client reported previous inconsistent use or the clinic staff suspected prior, recent ART use (i.e. a re-initiator presenting as naive). For the sake of completeness, we report results of these tests for participants who had them. Since the small number of participants who did receive baseline viral load tests is almost certainly non-representative of patients initiating or re-initiating ART, we did not include these data in the main analysis.

### Statistical analysis

Outcomes measured for each data source are shown in Table 1. We used these to create a variable to measure concordance between self-report and all other indicators of prior exposure. Those who self-reported no prior ART use and had no other positive indicators were designated “concordant-naïve.” Those who reported prior ART use but not in the past 90 days and had NHLS and/or EMR data that supported their timeline were labeled “concordant-experienced.” Finally, participants who self-reported no previous ART use but had evidence of prior ART experience by any other indicator were designated “discordant-experienced.”

We first summarized cohort and participant characteristics at ART initiation. Next, we arranged participants based on the three categories of concordance defined above and report results for individual participants to illustrate the range and frequency of each type of discordance. Then we combined all five measures of prior experience into a singular measure and used that as the gold standard to compare the sensitivity and negative predictive value of each of the individual measures. Because any singular positive indicator would deem a participant as treatment-experienced, false positives were not present. We therefore do not report specificity or positive predictive values, as these would equal 100% across all measures.

Interview transcripts were imported into NVivo. Interviews were read through then each question was coded line by line using a combination inductive-deductive approach and a content analysis was conducted. Key emerging themes for each question were summarized and supported by illustrative quotes which were lightly edited for clarity. Results are presented as integrated throughout the narrative using the contiguous approach[10].

### Ethics

The study protocol was reviewed and approved by the Human Research Ethics Committee of the University of Witwatersrand (M220440) and the Boston University IRB (H-42726) and by relevant Provincial Health Research Committees through the National Health Research Database in South Africa (KwaZulu-Natal: KZ_202207_014, Mpumalanga: MP_202207_004, Gauteng: GP_202207_049). The protocol is also registered on clinicaltrials.gov (NCT05454839). All participants provided written informed consent for all data collection.

## Results

### Study sample

Dried blood specimens were collected for 89 PREFER participants (median age 32.5, years, 62% female) (Table 2), of a total of 158 PREFER survey participants who met eligibility criteria for this sub-study. Of the 89 enrolled in the sub study between 29 August and 30 October 2023, 80 completed the full PREFER survey, while 9 completed an abbreviated survey. There were no important differences in terms of demographic and level of education, employment or whether they reported initiating ART for the first time, or were returning to care after a period of more than 90 days between the sample enrolled in the sub study and the full eligible population. Table 2 reports demographic and socioeconomic characteristics of the 80 who did complete the full survey.

**Table 2.** Characteristics of study participants who completed full PREFER questionnaire.

| Characteristic | N (%) |
| --- | --- |
| N | 80 |
| District* |  |
| West Rand, Gauteng Province | 21 (24%) |
| Ehlanzeni, Mpumalanga Province | 28 (31%) |
| King Cetshwayo, KZN Province | 40 (45%) |
| Gender (% female)* | 55 (62%) |
| Age* |  |
| Median (IQR) | 33 (27,38) |
| 18-24 years | 7 (8%) |
| 25-49 years | 73 (82%) |
| ≥50 years | 9 (10%) |
| Marital status |  |
| I have a primary partner or spouse who I live with | 25 (31%) |
| I have a primary partner or spouse but we do not live together | 41 (51%) |
| I do not have a primary partner or spouse at this time | 14 (18%) |
| How many adults and children are living in your household? |  |
| One person | 9 (11%) |
| 2-3 people adults | 28 (35%) |
| ≥4 people | 43 (54%) |
| How many other people in your household have HIV, to your knowledge? |  |
| None | 55 (69%) |
| One | 21 (26%) |
| 2 or more | 4 (5%) |
| Do you think of the house you currently live in as your main house? |  |
| Yes | 60 (75%) |
| No, my main house is somewhere else in South Africa | 14 (18%) |
| No, my main house is in another country | 6 (8%) |
| Number of years living in this area |  |
| <1 year | 14 (18%) |
| 1-4 years | 20 (25%) |
| 5-9 years | 46 (58%) |
| ≥10 years | 0 |
| Highest level of education |  |
| Primary or less | 35 (44%) |
| Secondary | 33 (41%) |
| Post-secondary | 12 (15%) |
| What is your primary occupation? |  |
| Formal employment | 30 (38%) |
| Informal employment | 9 (11%) |
| Unemployed | 39 (49%) |
| Student/trainee | 2 (3%) |
| Do you have electricity in your house? |  |
| No | 9 (11%) |
| Yes | 71 (89%) |
| Do you have access to piped water? |  |
| No | 3 (4%) |
| Yes – to house | 44 (55%) |
| Yes – community tap/pipe | 33 (41%) |
| How often do you or the people in your household go without food? |  |
| Never | 60 (75%) |
| Seldom | 6 (8%) |
| Sometimes | 13 (16%) |
| Often | 1 (1%) |
| If a person in your household became ill and 100 rands (approximately \$6) was needed for treatment, how difficult would it be for you to find this money? | |
| Difficult | 43 (54%) |
| Easy | 37 (46%) |

\*For the full sub-study sample of 89 participants; remaining data are only for n=80 who completed the full PREFER questionnaire.

### Indicators of prior exposure

Results for each indicator of prior ART exposure are shown in Table 3, with “X” designating no evidence of prior exposure and √ designating those with evidence of the specified exposure. Table 3 is ordered by concordance group (last column) as defined in the methods above.

**Table 3.**
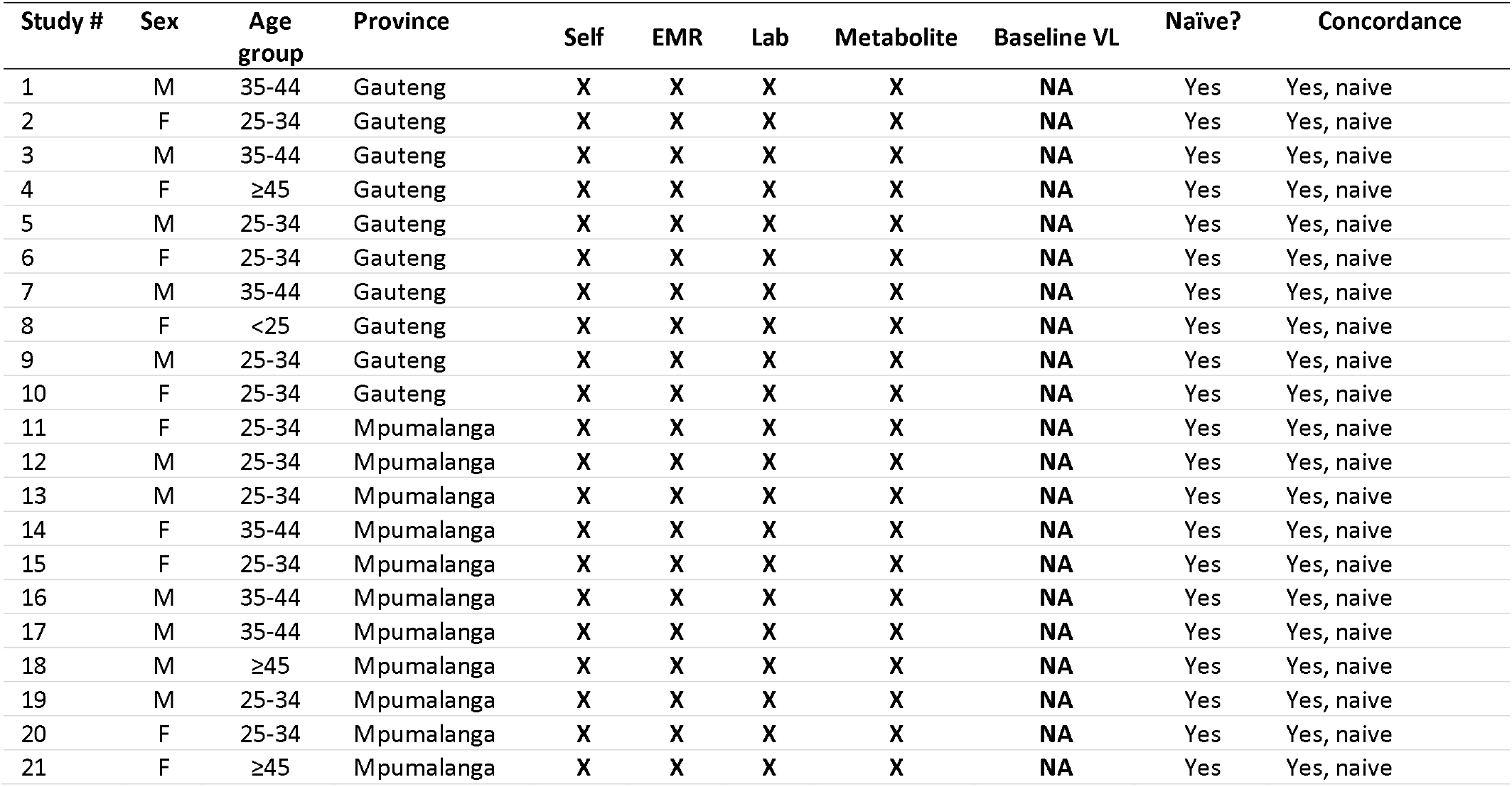
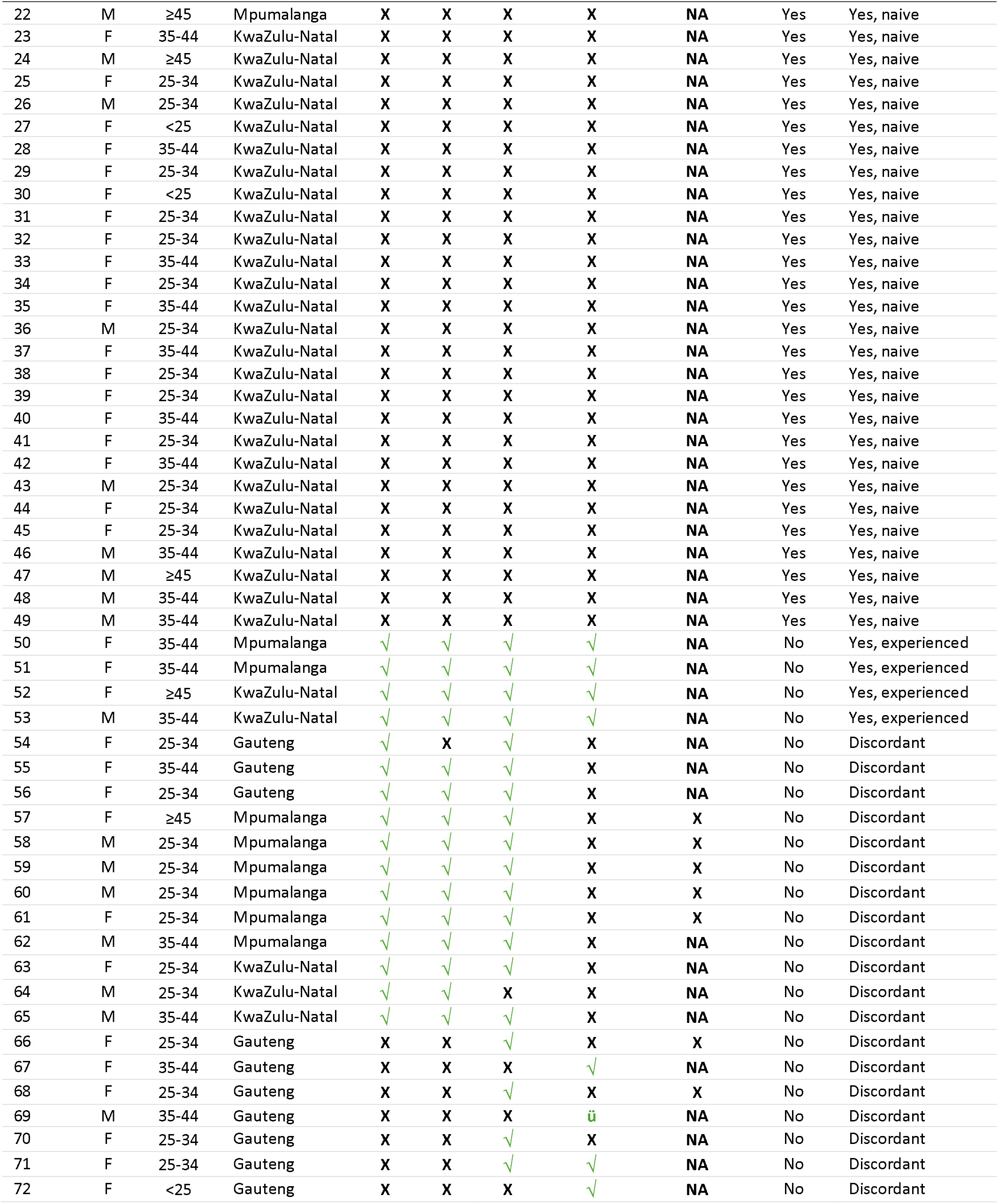
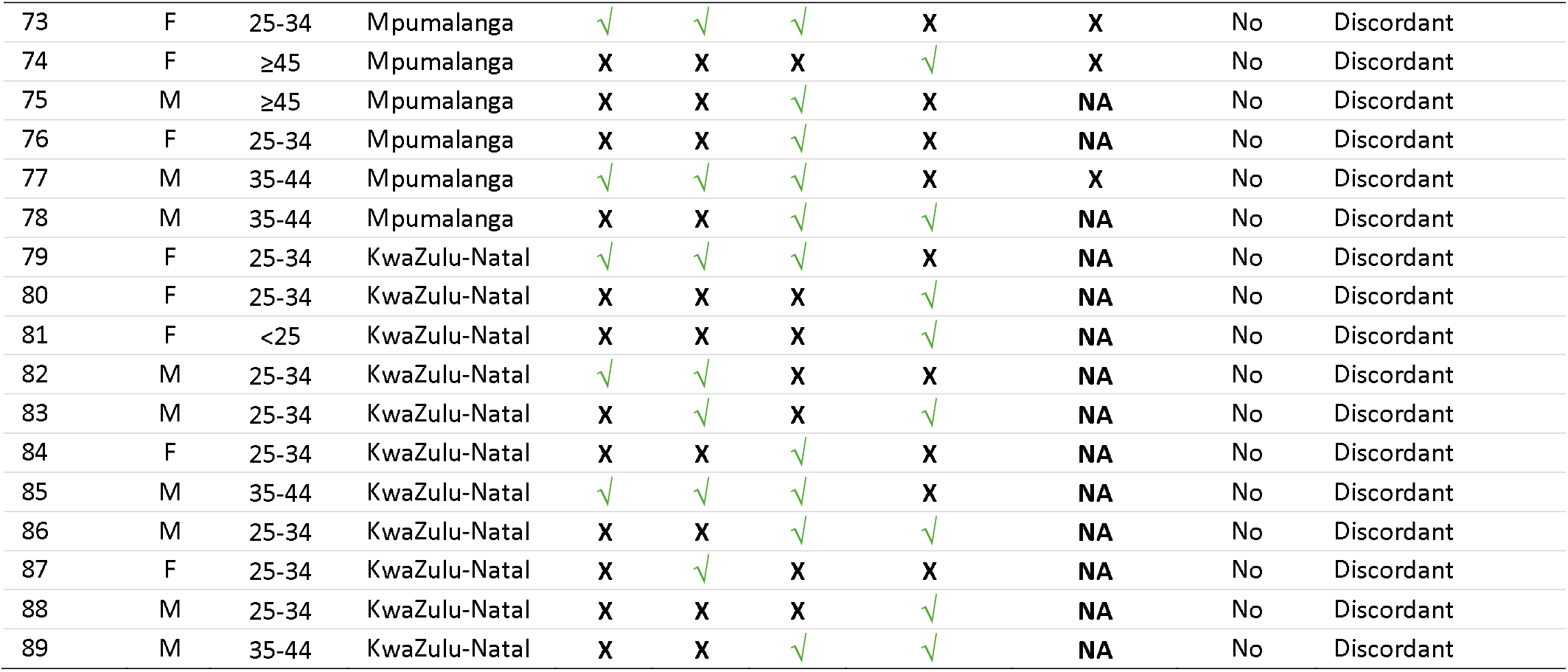
Exposure to prior ART for each study participant, by indicator.

Table 3 shows baseline viral loads conducted for 10 participants who received them. All baseline viral loads performed by the clinics were unsuppressed (i.e. did not indicate recent, previous ART exposure). As noted above, baseline viral load results were not considered in the analysis below.

Overall, 60% of participants (n=53) were concordant across all indicators. Most of these (n=49, 92% of those concordant, 55% of entire sample) were concordant-naive; 8% of those concordant and 45% of the full sample were treatment-experienced by at least one indicator. Only 4 participants had evidence of prior ART use across all the indicators (concordant-experienced). More than half (n=48, 53%) were discordant, with at least one but not all indicators showing prior ART use. Among the discordant, two thirds (n=24, 66% of discordant category) self-reported no prior exposure.

In Table 4, results for each indicator are aggregated to summarize study outcomes. For indicators that are not time limited, as shown in Table 1 (EMR records and lab records), we also report the proportion of those with record of prior ART use who also indicated prior use on the PREFER survey.

**Table 4.** Proportions showing prior ART exposure, by indicator (n=89)

| Indicator | N (%) |
| --- | --- |
| Self-report |  |
| No prior treatment experience | 73 (82%) |
| Prior treatment experience but no ART in past 90 days | 16 (18%) |
| Metabolite testing |  |
| No TDF detected | 71 (80%) |
| TDF detected | 18 (20%) |
| If detected, TDF level |  |
| <2 doses/week | 7 (8%) |
| 2-3 doses/week | 8 (9%) |
| 4-6 doses/week | 3 (3%) |
| EMR records |  |
| No evidence of prior ART | 72 (80%) |
| Evidence of ART >90 days before study enrollment | 17 (20%) |
| If evidence of ART >90 days before study enrollment, self-reported previous ART use on survey | 15 (88%) |
| Days between last evidence of ART care and current initiation (median, IQR) | 787 (333,1290) |
| Days since first ART initiation and last evidence of ART care prior to current initiation (median, IQR) | 456 (48,1260) |
| Laboratory records |  |
| No record of earlier viral load test (indication of prior ART use) | 70 (79%) |
| Record of viral load test in past, indicating ART use ≥90 days before study enrollment | 19 (21%) |
| If record of viral load test in past, self-reported previous ART use on survey | 10 (53%) |
| Composite result |  |
| Any indicator of prior ART use (experienced) | 49 (55%) |
| No indicator of prior ART use (naive) | 40 (45%) |

Most participants self-reported no prior ART experience at all on the survey (n=73, 82%). Sixteen (18%) self-reported previously taking ART but with a current treatment interruption of more than three months prior to presenting for ART re-initiation on the day of study enrollment.

Metabolite test results indicated that 18 (20%) participants had detectable TDF metabolites in their blood specimens. Among these 18, 3 (17%) had a level consistent with regularly taking 4-6 doses per week, 8 (44%) 2-3 doses per week, and 7 (38%) <2 doses per week. Of the 18 with detectable TDF metabolites, only four reported ever previously being exposed to ART, and all stated that the prior exposure was >90 days before the samples were taken. EMR and laboratory records both indicated that about one fifth of participants had evidence of previous ART; 88% of those with EMR records and 53% of those with lab evidence disclosed this prior use when responding to the PREFER survey. In the EMR, the median time since last evidence of ART treatment--the duration of the interruption as documented in the EMR--was just over two years (787 days). The median time in care between first ART initiation and last evidence of ART care--time on ART prior to interruption--was roughly one and quarter years (456 days).

### Accuracy of indicators

To compare the accuracy of each indicator in identifying prior ART exposure, we considered the composite measure of treatment experience reported in Table 4, using all five potential indicators of prior exposure, as our gold standard to calculate the sensitivity and negative predictive value of each method (Table 5). Evidence of prior treatment in laboratory records was the most sensitive (73%) and had the highest negative predictive value (82%), while self-report fared the worst (sensitivity 40%, NPV 67%). Metabolite testing and EMR evidence had similar sensitivities and NPRs. Combining indicators had mixed results for increasing accuracy, due to the varying degree of overlap between positive indicators. The sensitivity of self-report plus laboratory record was markedly higher than self-report alone (75% vs 40%) but was not a substantial improvement over the laboratory record on its own (73%). Of all combinations, laboratory record of prior VL plus TDF metabolite was the most accurate, with a sensitivity of 95% and NPV and 96%.

**Table 5:** Sensitivity and negative predictive value of measures in detecting prior treatment exposure.

| Method | Sensitivity | Negative predictive value |
| --- | --- | --- |
| Self-report | 40% | 67% |
| Laboratory record of prior VL | 73% | 82% |
| EMR prior evidence | 43% | 68% |
| TDF metabolite | 45% | 69% |
| Self-report + Laboratory record of prior VL | 75% | 83% |
| Self-report + EMR prior evidence | 45% | 69% |
| Self-report + TDF metabolite | 75% | 83% |
| Laboratory record of prior VL + EMR prior evidence | 80% | 86% |
| Laboratory record of prior VL + TDF metabolite | 95% | 96% |
| EMR prior evidence + TDF metabolite | 75% | 83% |

### Predictors of prior exposure

Neither gender nor age was associated with having evidence of prior treatment experience (Supplementary Table 1). Those with prior treatment experience were more likely to have primary schooling or less (52%) than those who were naïve (36%) and were less likely to be formally employed (24% vs 50%) and more likely to be unemployed (55% vs 43%) than naïve participants. There were no substantial differences in access to utilities, food security, or ability to pay for medical expenses between treatment experience groups. Patients attending the study facility in the King Cetshwayo District were less likely to have prior exposure (34%) than were those from Ehlanzeni District (59%) or West Rand District (52%).

### Reasons for non-disclosure

Between 30 April and 8 July 2024, qualitative interviews were conducted with 11 of the 18 participants who self-reported no prior ART exposure but had detectable TDF metabolites in their blood specimens. We were unable to contact the remaining seven despite several attempts. Of those contacted, two were from West Rand District, three from Ehlanzeni District, and six from King Cetshwayo District. Six of the 11 were female.

Primary responses to the discordant results (self-report v metabolite results) included use of pre-exposure prophylaxis (PrEP) (n=1), denial of prior use despite the metabolite evidence (n=3), and, most commonly, admission of prior use and an explanation that presenting as naive is preferable for the patient (n=7). One respondent explained that she had taken PrEP but had stopped several months prior to testing HIV-positive. She attended the clinic to obtain a pregnancy test and received a routine HIV test as part of antenatal care. She enrolled in the study and provided a blood sample for metabolite testing the same day. Three of the participants continued to deny any known exposure to ART, as illustrated by this quote: *“I have never taken treatment before that day, I am sure about that and I was telling truth*.*” (*Female, West Rand District.)

While each of the remaining seven respondents had a unique explanation, all intentionally presented as new (naive) clients on the day of enrollment.

One participant implied that the clinic staff had encouraged him to start as a new patient. *“When I relocated to [Clinic X], I went to the clinic and they told me to start afresh as I am like a person who has never taken treatment before, they tested me that day and started everything from scratch*.*”* (Male, King Cetshwayo District.)

Three participants had personal circumstances or experiences that made it physically or emotionally challenging to continue ART.

- One had stopped taking treatment because an injury made it difficult to get to the clinic. *“I started taking treatment in 2016, I got seriously injured and where I was staying at that time it was very difficult for me to go to the clinic for treatment hence I ended stop taking them*.*”* (Male, Ehlanzeni District.)
- Another implied that her partner was not supportive, which she found stressful and logistically difficult, though it is unclear from her interview if she had actually stopped. *“I was feeling so sad that day and I was scared to disclose a lot during that day [referencing both her partner and ART status]. I think it is stress caused by my partner. He doesn’t want me to take treatment so I end up hiding my treatment in the house*.*”* (Female, Ehlanzeni District)
- The third described a negative interaction with the provider, causing her to stop treatment. *“I [had] started treatment in 2017, but I stopped taking them for [unknown period of time], because I had an issue with the nurse. I had skipped my next scheduled appointment, and when I went back to the clinic the nurse mistreated me so I decided to stop going to the clinic. I was in the queue the entire day and when I was in front of the queue, then she told me because I had missed my date I will have to go to the back she will attend me at last and I had asked at work so I was not treated well then, I stopped*.*”* (Female, King Cetshwayo District)

Finally, three respondents described that they travel frequently for work and explained that they find it simpler and easier to either share medication or present as new patients, rather than deal with the consequences of transferring, which they described as including being shouted at by providers and needing transfer paperwork that can be difficult to obtain.

- *“The issue is I’m someone who is travelling quite a lot, so sometimes when my clinic date comes and I am not around, I end up sharing treatment with the mother of my children, I would appear as someone who has defaulted on treatment at the clinic yet I am using it from the mother of my children. So, when I go back to the clinic after a long time appeared as defaulted treatment and I just present as a new patient*.*”* (Male, King Cetshwayo District)
- *“It just that the issue of job opportunities I move around a lot, so wherever I am at that point in time*
- *when I need treatment I go to the nearest clinic where I present myself as a new patient to avoid delays and asked a lot of questions. So, in order for me to access treatment easily without being shouted at or asked many questions or required documents such as transfer letters from previous clinics that may lead me not to get treatment, I just test then start treatment. So, this becomes an easy way to get treatment*.*”* (Male, King Cetshwayo District)
- *“When those ladies asked me during the interview about my treatment history I said no because I have never officially started treatment but I have taken treatment through my partner whenever she was around [and supervisor], so I was scared to say it because I have never officially gone and start treatment at the clinic*.*”* (Male, King Cetshwayo District.)

When asked about how facility staff could make it easier for patients to share their prior treatment history, emerging themes in interview responses included improving privacy, improving tolerance and patience among nurses, increasing counselling and outreach with peer ambassadors, and including clients in the decision-making process, as illustrated below.

- *“I think nurses should ask questions about our treatment journeys not tell us, including when they have to give us next appointment dates they need to ask us how comfortable are we with such dates are we going to be able to come to the facility and we share our information or experiences with them, not being told on what to do and disregards our life situations*.*”* (Male, Ehlanzeni District.)

Five respondents offered thoughts on why other people in the community may not remember or disclose that they have had prior ART exposure. Among emerging themes were that patients truly forget because they are now feeling better; patients perceive stigma from health facility staff for having defaulted; and that it is simpler to start as a new patient than to present as a re-initiator.

- *“If I guess, maybe when someone feels better in his or her body may end up forgetting that they have taken treatment before or even remembering when asked because their body is functioning well there are no signs of sickness*.*”* (Male, King Cetshwayo District.)
- *“Yes, [presenting new] is something that happens, as I did myself and it works for me imagine how many people in the world taking treatment, don’t you think there are others doing the same thing? Although I can’t say I know someone specifically but for sure there are many people doing this*.*”* (Male, King Cetshwayo District.)
- *“Maybe [they are] scared just like myself I was scared. Most of the time in clinics they shout at us if you have defaulted or missed appointments. They will shout you so bad making us scared, hence they just say they have never taken treatment, they know they have but they scared of attitude and treatment they will get that is why they end up not giving correct information or say they never taken treatment before*.*”* (Male, Ehlanzeni District.)
- *“There are a lot of people saying they have never taken treatment when they have. Sometimes us as guys we go to check and after checking they will say we must start treatment and for whatever reason we don’t. Then we go elsewhere to want to start treatment, to avoid a long process and a lot of explanation in new facilities people just decide to say they are new on treatment to cut off a lot of questions and challenges to access treatment*.*”* (Male, King Cetshwayo District.)

## Discussion

In this mixed-methods study of a sequential sample of adults presenting to initiate or re-initiate HIV treatment in South Africa and self-reporting no prior ART use within the past 90 days, 55% had at least one indicator suggesting previous ART use. Roughly one fifth were found to have ARV metabolites in blood samples indicating recent (within 90-day) use. An additional third had records of prior viral load tests or ART care, evidence that they were likely restarting ART after a short or long interruption. The qualitative interviews of 11 respondents with discordant results suggest a variety of reasons for both interrupting treatment and non-disclosure of prior experience, including deliberately presenting as new to avoid potential stigma from clinic staff and burdensome transfer processes.

The primary strength of this study was the use of multiple data sources to find evidence of prior ART exposure in the same sample. We found a high level of non-concordance among the sources, with only 10% of those who appear to have prior ART experience showing positive indicators across all sources. This is not entirely surprising, as some indicators—self report and EMR and lab records—represent lifetime experience, while others—metabolite testing (and baseline viral loads, to the extent captured)—reflect only recent use. It does make clear that none of the available indicators alone is sufficient for an accurate estimate. In our study, laboratory records and EMR records were the most accurate, while self-report was the least.

Metabolite testing and baseline viral load tests, both of which can accurately discern recent ART use, also both pose a number of problems for routine use. Metabolite testing is expensive, can only be carried out in specialized laboratories and, in our analysis, did not perform better than self-report. Baseline viral load tests, which are being introduced into routine care in some settings[11] but are not recommended as part of routine care in South African treatment guidelines, are less expensive but, in the absence of rapid point-of-care technologies, provide information only days or weeks after the initiation visit, by which time many patients have already disengaged from care. Both of these approaches, moreover, only capture ART exposure within the past 1-3 months, before viral rebound or metabolite clearance. In view of this, active review of patients’ clinic records (EMR) and/or laboratory records may be the most practical approach to identifying re-initiators, particularly where such records are available to clinicians in real time at point of care. In South Africa, for example, a counselor or clerk may be able to look up patients’ clinic and lab histories while a patient waits for services and convey any evidence of prior treatment to the nurse before the treatment initiation consultation begins.

This study follows on a systematic review we conducted that found limited data on prior treatment exposure[12]. We found that there was no consistency in defining populations where prior treatment exposure was being reported and varied estimates in prior exposure measured in similar ways. For example, self-reported estimates in the literature range from 2-21%[13–18], which places our estimate of 18% on the high end. When using EMR data, estimates have ranged from 22% in a population that had ART naivete in the inclusion criteria[19] to 69% in a dataset that considered all initiations and allowed for individuals to be tracked across multiple facilities[3]. Metabolite testing also ranged from 19% when testing only blood specimens for tenofovir and emtricitabine among participants with undetectable viral loads at initiation to 53% when using both blood and hair samples to test for tenofovir, efavirenz and emtricitabine among a study population who all reported being ART naive[19,20]. Baseline viral load suppression is another measure reported in the literature with relatively high yield of previous exposure (30-52%)[21,22]. Among the small number of participants in our study who had a viral load performed as part of their initiation laboratory tests, however, none were suppressed.

A further strength of this study is that in addition to these measures, we also conducted qualitative interviews with participants who had tested positive for ART metabolites after reporting being treatment naïve or not using ART in the preceding 90 days. One interviewee reported using PrEP, which commonly includes tenofovir and can remain detectable in dried blood specimens for up to 3 months[9]. As PrEP use expands, testing for metabolites of drugs common to PrEP and ART may become less useful. It is perplexing and concerning that three of the 11 participants we interviewed continued to deny any known prior exposure to ART before the day of enrollment into the study. While it is possible that participants did not remember prior use or were inadvertently exposed through recreational drugs, neither of these explanations seems likely. We speculate that it is more likely that these three individuals simply represent more adamant examples of the seven clients who deliberately presented to the facilities as naive and determined that it was in their interest not to reveal prior exposure even to researchers.

For those who did reveal to our interviewer that they had deliberately concealed their prior ART use, the quotations included here offer a range of reasons that are consistent with prior research[23,24] and appear eminently rational from the perspective of the patients. Despite national guidelines that advise clinic staff to be welcoming and encouraging to re-engagers[25–27], there is abundant evidence that facilities and providers deter patients from revealing prior disengagement[23,24,28]. Previous research suggests that provider disposition, in particular, is an important driver of patient behavior [29]. Re-engaging patients report being chastised (or worse), sent to the back of the queue, or told to return on another day with an official transfer letter from another facility. All of these actions on the part of providers would lead patients to hide their previous treatment, particularly when initiation procedures for naive patients have become simpler and faster in recent years [30,31]. The time required to locate existing files is also an obstacle, compared to opening a new file. It is striking that one respondent reported that clinic staff advised him to present as a new patient, suggesting that staff, too, regard the re-engagement process as burdensome.

Our study had a number of limitations, leading us to regard it as exploratory rather than definitive. First, our inclusion criteria required participants to conform that they were ART naïve or had a treatment interruption of 90 days or more and were informed that we would conduct metabolite testing and search medical records for prior ART exposure. This process may have led some potential participants to self-select out of the study at the time of screening because they were aware of having undisclosed, prior treatment experience, leading us to underestimate the proportion of initiators with positive measures of prior exposure. Second, we could not access EMR records from other clinics participants may have attended previously, making our EMR-based result for prior exposure almost certainly an under-estimate. Third, we did not have baseline viral load test results for most participants. We did conduct viral load tests on the same dried blood specimens used for metabolite testing for all participants, but results were sufficiently inconsistent with the routinely reported viral load tests conducted by clinics for the 10 clients mentioned above that we discarded the study DBS findings as unreliable. This may have been due to a long delay between sample collection and ultimate viral load testing of DBS or the unreliability of DBS samples for viral load estimates. Fourth, while we are not aware of other studies using multiple data sources in the same sample to compare the sensitivity of different indicators, our sample was small and limits our ability to comment on predictors of prior treatment exposure with confidence. Fifth, our small number of facilities in just three districts makes generalizability to other locations uncertain, such that our results should be regarded as exploratory only. Sixth, there may be a difference between self-reported prior exposure reported to study interviews and that revealed to clinic staff. Finally, our qualitative analysis might have been subject to survey bias and we are uncertain if we reached saturation or predictability, though we did see repeated themes that elicited important nuance. Despite these limitations, our results are consistent with prior studies using single indicators.

## Conclusions

In this exploratory study, we found evidence that more than half of adults who present for ART initiation in South Africa have previous treatment exposure, although most prefer not to reveal that experience to healthcare providers due to the negative attitude that the healthcare providers portray towards them. To the extent that knowledge of a patient’s prior experience on ART is an important indicator of future outcomes and can guide providers toward effective interventions, these results pose a major challenge to improving the quality of HIV care. These results could further guide and improve the review of the National Welcome Back Strategy. Future work should explore reasons for reluctance to disclose prior ART use, how to identify re-engagers at initiation, interventions needed to support re-engagers, and healthcare providers’ reasons for not prioritizing the re-engagers in care. In the meantime, providers would be justified in assuming that prior ART experience is the norm, not the exception, when patients present for ART initiation, and offer information, support, and services accordingly.

## Data Availability

Disidentified data generated by the study (questionnaire responses, biological sample results) will be made available in a public data repository within one year of study protocol closure by the responsible ethics committees. Data obtained from participants routinely collected clinic and laboratory records are owned by the South Africa National Department of Health and cannot be shared by the authors.

## Supporting information

Supplementary File 1: Creation of DBS (dried blood specimen) samples for metabolite testing

Supplementary Table 1: Participant characteristics by prior exposure status for participants who completed the full PREFER survey

## Acknowledgements

None.

## Authors’ contributions

MM, MB, and SR conceived of and designed the study. MB, NS, and VN developed the qualitative instrument. MB conducted the quantitative data analysis with input from MM, SR, and BN. VN and NN conducted, translated, and transcribed the qualitative interviews. NS analyzed the qualitative data. MB, MM, SR, and NS drafted the initial manuscript. LM and MM interpreted the results. All authors reviewed the manuscript and approved the final version.

## Registration

The protocol is registered on clinicaltrials.gov (NCT05454839).

