## Supplementary file 1 for "Prior antiretroviral therapy exposure among clients presenting for HIV treatment initiation in South Africa: an exploratory mixed-methods study using multiple indicators of exposure"

Supplementary file 1: Creation of DBS (dried blood specimen) samples for metabolite testing

DBS samples consisting of 5 x 50μml dried blood spots on a Whatman 903 Protein saver card were created from venous specimens collected into an EDTA blood tube at ART initiation at the time of treatment initiation. Capillary tubes were initially used to measure 50μml per spot, but after quality concerns were raised, they were replaced with one, single use, 50μml pipette.

Specimens were allowed to air dry for a minimum of 2 hours before being closed using the flap included on the card and placed in a non-gas permeable bag with a humidity indicator card and desiccant sachet. Bags were placed in a clearly marked polystyrene box in a freezer (-4 degrees Celsius). Both the freezer temperature and humidity indicators were checked on a weekly basis. Desiccant packs were replaced if the humidity indicator was positive. No samples were kept in freezers prior to shipping to the analyzing lab for longer than 2 months.

Specimen cards were marked with participants’ study ID and date and time of collection prior to creation of the specimen using a permanent marker. The study ID, date and time of collection and participant initials were captured in a sample log. Sample log and specimen numbers were reconciled both prior to shipping specimens from the facilities and on arrival at the laboratory in Cape Town.
