## Supplementary table 1 for "Prior antiretroviral therapy exposure among clients presenting for HIV treatment initiation in South Africa: an exploratory mixed-methods study using multiple indicators of exposure"

**Supplementary Table 1: Participant characteristics by prior exposure status for participants who completed the full PREFER survey**

| **Characteristics** | **≥1 indicator of prior exposure** | **No prior exposure** | **Overall** |
| --- | --- | --- | --- |
| N | 38 | 42 | 80 |
| District |  |  |  |
| West Rand | 11 (28.9%) | 10 (23.8%) | 21 (26.3%) |
| Ehlanzeni | 16 (42.1%) | 11 (26.2%) | 27 (33.8%) |
| King Cetshwayo | 11 (28.9%) | 21 (50.0%) | 32 (40.0%) |
| Age |  |  |  |
| Median [IQR] | 32.0 [27, 38] | 33.0 [28, 40] | 33.0 [27, 38] |
| 18-25 | 4 (10.0%) | 3 (6.1%) | 7 (7.9%) |
| 25-49 | 32 (80.0%) | 41 (83.7%) | 73 (82.0%) |
| 50+ | 4 (10.0%) | 5 (10.2%) | 9 (10.1%) |
| Sex (female) | 24 (63.2%) | 26 (61.9%) | 50 (62.5%) |
| Highest level of education |  |  |  |
| Primary or less | 20 (52.6%) | 15 (35.7%) | 35 (43.8%) |
| Secondary | 16 (42.1%) | 17 (40.5%) | 33 (41.3%) |
| Post-secondary | 2 (5.3%) | 10 (23.8%) | 12 (15.0%) |
| Marital status |  |  |  |
| I have a primary partner or spouse who I live with | 11 (28.9%) | 14 (33.3%) | 25 (31.3%) |
| I have a primary partner or spouse but we do not live together | 19 (50.0%) | 22 (52.4%) | 41 (51.3%) |
| I do not have a primary partner or spouse at this time | 8 (21.1%) | 6 (14.3%) | 14 (17.5%) |
| How many living children do you have? | | | |
| No children | 8 (21.1%) | 12 (28.6%) | 20 (25.0%) |
| 1-2 children | 17 (44.7%) | 19 (45.2%) | 36 (45.0%) |
| 3 or more children | 13 (34.2%) | 11 (26.2%) | 24 (30.0%) |
| How many adults and children are living in your household? | | | |
| 1 | 5 (13.2%) | 4 (9.5%) | 9 (11.3%) |
| 2-3 | 11 (28.9%) | 17 (40.5%) | 28 (35.0%) |
| ≥4 | 22 (57.9%) | 21 (50.0%) | 43 (53.8%) |
| How many other people in your household have HIV, to your knowledge? | | | |
| None | 25 (65.8%) | 30 (71.4%) | 55 (68.8%) |
| 1 | 10 (26.3%) | 11 (26.2%) | 21 (26.3%) |
| ≥2 | 3 (7.9%) | 1 (2.4%) | 4 (5.0%) |
| Do you think of the house you currently live in as your main house? | | | |
| Yes | 28 (73.7%) | 32 (76.2%) | 60 (75.0%) |
| No, my main house is somewhere else in South Africa | 7 (18.4%) | 7 (16.7%) | 14 (17.5%) |
| No, my main house is in another country | 3 (7.9%) | 3 (7.1%) | 6 (7.5%) |
| Number of years living in this area |  |  |  |
| Less than a year | 8 (21.1%) | 6 (14.3%) | 14 (17.5%) |
| 1-4 years | 9 (23.7%) | 11 (26.2%) | 20 (25.0%) |
| 5-9 years | 21 (55.3%) | 25 (59.5%) | 46 (57.5%) |
| What is your reading level? |  |  |  |
| Read well | 30 (78.9%) | 37 (88.1%) | 67 (83.8%) |
| Read somewhat | 8 (21.1%) | 5 (11.9%) | 13 (16.3%) |
| How comfortable are you speaking English? | | | |
| I am very comfortable | 27 (71.1%) | 36 (85.7%) | 63 (78.8%) |
| I am somewhat comfortable | 11 (28.9%) | 6 (14.3%) | 17 (21.3%) |
| How comfortable are you using a mobile phone or computer for receiving information | | | |
| I am very comfortable | 27 (71.1%) | 36 (85.7%) | 63 (78.8%) |
| I am somewhat comfortable | 11 (28.9%) | 6 (14.3%) | 17 (21.3%) |
| What is your primary occupation? |  |  |  |
| Formal employment | 9 (23.7%) | 21 (50.0%) | 30 (37.5%) |
| Informal employment | 6 (15.8%) | 3 (7.1%) | 9 (11.3%) |
| Unemployed | 21 (55.3%) | 18 (42.9%) | 39 (48.8%) |
| Student/Trainee | 2 (5.3%) | 0 (0%) | 2 (2.5%) |
| What time of day do you usually work? | | | |
| All day (regular working day) | 5 (13.2%) | 19 (45.2%) | 24 (30.0%) |
| Afternoons only | 1 (2.6%) | 0 (0%) | 1 (1.3%) |
| Flexible times (work for myself) | 2 (5.3%) | 2 (4.8%) | 4 (5.0%) |
| Shifts change from day to day | 8 (21.1%) | 3 (7.1%) | 11 (13.8%) |
| I do not work | 22 (57.9%) | 18 (42.9%) | 40 (50.0%) |
| Do you have electricity in your house? | | | |
| No | 3 (7.9%) | 6 (14.3%) | 9 (11.3%) |
| Yes | 35 (92.1%) | 36 (85.7%) | 71 (88.8%) |
| Do you have access to piped water? | | | |
| No | 2 (5.3%) | 1 (2.4%) | 3 (3.8%) |
| Yes – to house | 19 (50.0%) | 25 (59.5%) | 44 (55.0%) |
| Yes – community tap/pipe | 17 (44.7%) | 16 (38.1%) | 33 (41.3%) |
| Do you or the people in your household go without food often, sometimes, seldom? | | | |
| Never | 28 (73.7%) | 32 (76.2%) | 60 (75.0%) |
| Seldom | 3 (7.9%) | 3 (7.1%) | 6 (7.5%) |
| Sometimes | 7 (18.4%) | 6 (14.3%) | 13 (16.3%) |
| Often | 0 (0%) | 1 (2.4%) | 1 (1.3%) |
| If a person in your household became ill and 100 Rands was needed for treatment, how difficult would it be for you to get it? | | | |
| Difficult | 23 (53%) | 21 (47%) | 43 (54%) |
| Easy | 16 (43%) | 21 (57%) | 37 (46%) |
